# Will COVID-19 pandemic diminish by summer-monsoon in India? Lesson from the first lockdown

**DOI:** 10.1101/2020.04.22.20075499

**Authors:** Sarvan Kumar

## Abstract

The novel Coronavirus (2019-nCoV) was identified in Wuhan, Hubei Province, China, in December 2019 and has created a medical emergency worldwide. It has spread rapidly to multiple countries and has been declared a pandemic by the World Health Organization. In India, it is already reported more than 18 thousand cases and more than 600 deaths due to Coronavirus disease 2019 (COVID-19) till April 20, 2020. Previous studies on various viral infections like influenza have supported an epidemiological hypothesis that the cold and dry (low absolute humidity) environments favor the survival and spread of droplet-mediated viral diseases. These viral transmissions found attenuated in warm and humid (high absolute humidity) environments. However, the role of temperature, humidity, and absolute humidity in the transmission of COVID-19 has not yet been well established. Therefore the study to investigate the meteorological condition for incidence and spread of COVID-19 infection, to predict the epidemiology of the infectious disease, and to provide a scientific basis for prevention and control measures against the new disease is required for India. In this work, we analyze the local weather patterns of the Indian region affected by the COVID-19 virus for March and April months, 2020. We have investigated the effect of meteorological parameters like Temperature, relative humidity, and absolute humidity on the rate of spread of COVID-19 using daily confirm cases in India. We have used daily averaged meteorological data for the last three years (2017-2019) for March and April month and the same for the year 2020 for March 1 to April 15. We found a positive association (Pearson’s r=0.56) between temperature and daily COVID-19 cases over India. We found a negative association of humidity (RH and AH) with daily COVID-19 Cases (Person’s r=-0.62, -0.37). We have also investigated the role of aerosol in spreading the pandemic across India because it’s possible airborne nature. For this, we have investigated the association of aerosols (AOD) and other pollutions (NO_2_) with COVID-19 cases during the study period and also during the first lockdown period (25 March-15 April) in India. We found a negative association in March when there were few cases, but in April, it shows positive association when the number of cases is more (for AOD it was r=-0.41 and r=0.28 respectively). During the lockdown period, aerosols (AOD) and other pollutants (NO_2_; an indicator of PM_2.5_) reduced sharply with a percentage drop of about 36 and 37, respectively. This reduction may have reduced the risk for COVID-19 through air transmission due to the unavailability of aerosol particles as a base. HYSPLIT forward trajectory model also shows that surface aerosols may travel up to 4 km according to wind and direction within three h of its generation. If coronavirus becomes airborne as suggested by many studies, then it may have a higher risk of transmission by aerosols particles. So relaxing in the lockdown and environmental rules in terms of pollutant emissions from power plants, factories, and other facilities would be a wrong choice and could result in more COVID-19 incidences and deaths in India. Therefore the current study, although limited, suggests that it is doubtful that the spread of COVID-19 would slow down in India due to meteorological factors, like high temperature and high humidity. Because a large number of cases have already been reported in the range of high Tem, high Relative, and high absolute humidity regions of India. Thus our results in no way suggest that COVID-19 would not spread in warm, humid regions or during summer/monsoon. So effective public health interventions should be implemented across India to slow down the transmission of COVID-19. If COVID-19 is indeed sensitive to environmental factors, it could be tested in the coming summer-monsoon for India. So the only summer is not going to help India until monsoon is coming. Only government mitigations strategies would be helpful, whether its lockdown, aggressive and strategic testing, medical facilities, imposing social distancing, encouraging to use face mask or monitoring by a mobile application (Aarogya Setu).

**Highlights:**

- First study on the effects of meteorological factors on COVID-19 cases in India.
- A positive association between daily new cases of COVID-19 with temperature.
- RH and AH are negatively associated with daily new cases of COVID-19.
- Early lockdown in India slows down the spread of contagious disease COVID-19.
- More than a 35% fall was found in AOD and NO_2_ values during the lockdown period.

**Graphical abstract:** Correlation of daily confirmed cases of COVID-19 with the Temperature, Relative Humidity, and Absolute humidity from 20 March -15 April 2020 for the Indian region.

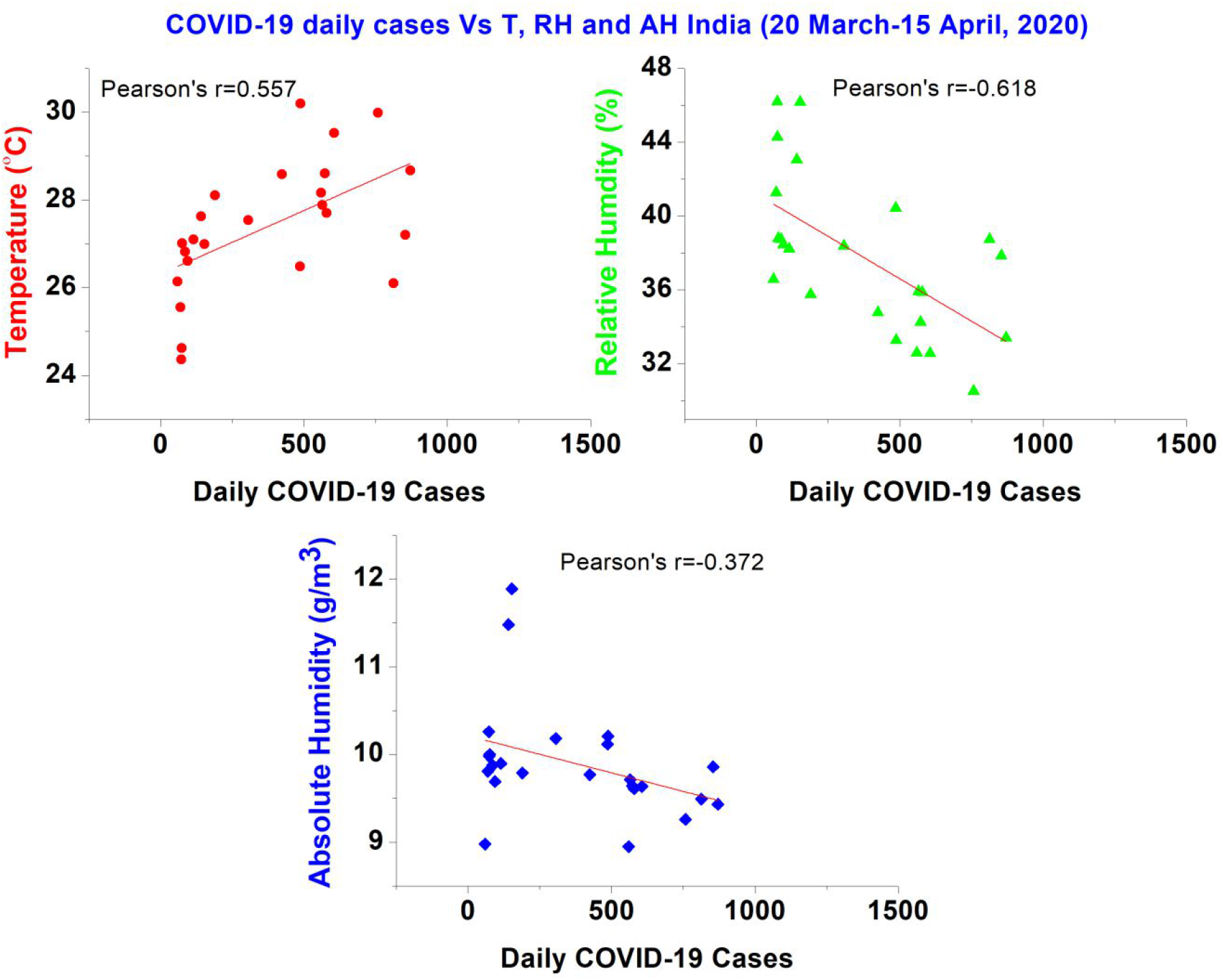

## 1. Introduction

The novel Coronavirus (2019-nCoV) was identified in Wuhan, Hubei Province, China, in December 2019 (Bukhari and Jameel, 2020) and caused over 2.46 million cases and over 170 thousand deaths worldwide till date (20 April 2020) (Worldometer). It has spread rapidly to multiple countries and has been declared a pandemic by the World Health Organization on March 11, 2020 (WHO). In India, it is already reported more than 18 thousand cases and more than 600 deaths due to Coronavirus disease 2019 (COVID-19) (COVID/Tracker). Previous studies have supported an epidemiological hypothesis that cold and dry (low absolute humidity) environments facilitate the survival and spread of droplet-mediated viral diseases. Warm and humid (high absolute humidity) environments see attenuated viral transmission like influenza and SARS (Schoeman and Fielding, 2019). As this coronavirus appeared for the first time and was highly contagious, it poses a great challenge to diagnosis and prevention and control. Human coronaviruses have been associated with a wide spectrum of respiratory diseases in different studies and belong to the *Coronaviridae* family (Bukhari and Jameel, 2020; Weiss and Navas-Martin, 2005). It has been suggested that flu viruses are not easily transmitted in hot and humid conditions. Similar comments about the COVID-19 have repeatedly been made by health officials as well as world leaders that the outbreak will slow down by summer, due to decreased transmissivity (Bukhari and Jameel, 2020). Wang et al. (2020) also found a similar result in his model study. They suggested that during the coming summer in the northern hemisphere, the spread of Coronavirus will be reduced in tropical regions. It is also important to note that SARS-Cov, which is a type of coronavirus, loses its ability to survive in higher temperatures, which may be due to the breakdown of their lipid layer at higher temperatures (Schoeman and Fielding, 2019). However, no seasonality has been established for COVID-19.

Bukhari and Jameel (2020) reported that in the beginning none of the Asian, Middle Eastern and South American countries had implemented drastic quarantine measures such as those in China, Europe, and some US states, however, their overall growth rate was lower, but now the rate is much similar to the Europe and USA. He suggested that it could be due to a lower number of testing, such as in India, Pakistan, Indonesia, and African countries. Many countries such as Singapore, UAE, Saudi Arabia, Australia, Qatar, Taiwan, and Hong Kong have performed more 2019-nCoV tests per million people than the USA, Italy, and several European countries. It was suggested that non-testing was not an issue, at least for the tropical countries.

At the beginning of April, thousands of new cases have been documented in regions with Tem >18 C, suggesting that the role of warmer temperature in slowing the spread of the COVID-19, as suggested earlier, might only be observed, at much higher temperatures. Unlike temperature, most of the COVID-19 cases were reported in the range of AH has consistently been between 3 and 9 g/m^3^ (Bukhari and Jameel, 2020). Bukhari and Jameel, (2020) also suggested that if, new cases in April and May continue to cluster within the observed range of AH, i.e., 3 to 9g/m^3^, then the countries experiencing monsoon, i.e., having high absolute humidity (>10 g/m^3^) may see a slowdown in transmissions, due to climatic factors. But for India, it is not true as many states having high temperate and high humidity are still leading in COVID-19 cases in India like Maharashtra and Tamil Nadu (Fig. 1) as for India, the average AH, is between 8 to 11 g/m^3^ during March and April month. A higher number of cases also reported for Kerala and Uttar Pradesh at the beginning of April, but government early mitigation strategies controlled the daily new COVID-19 cases.

**Fig 1.**
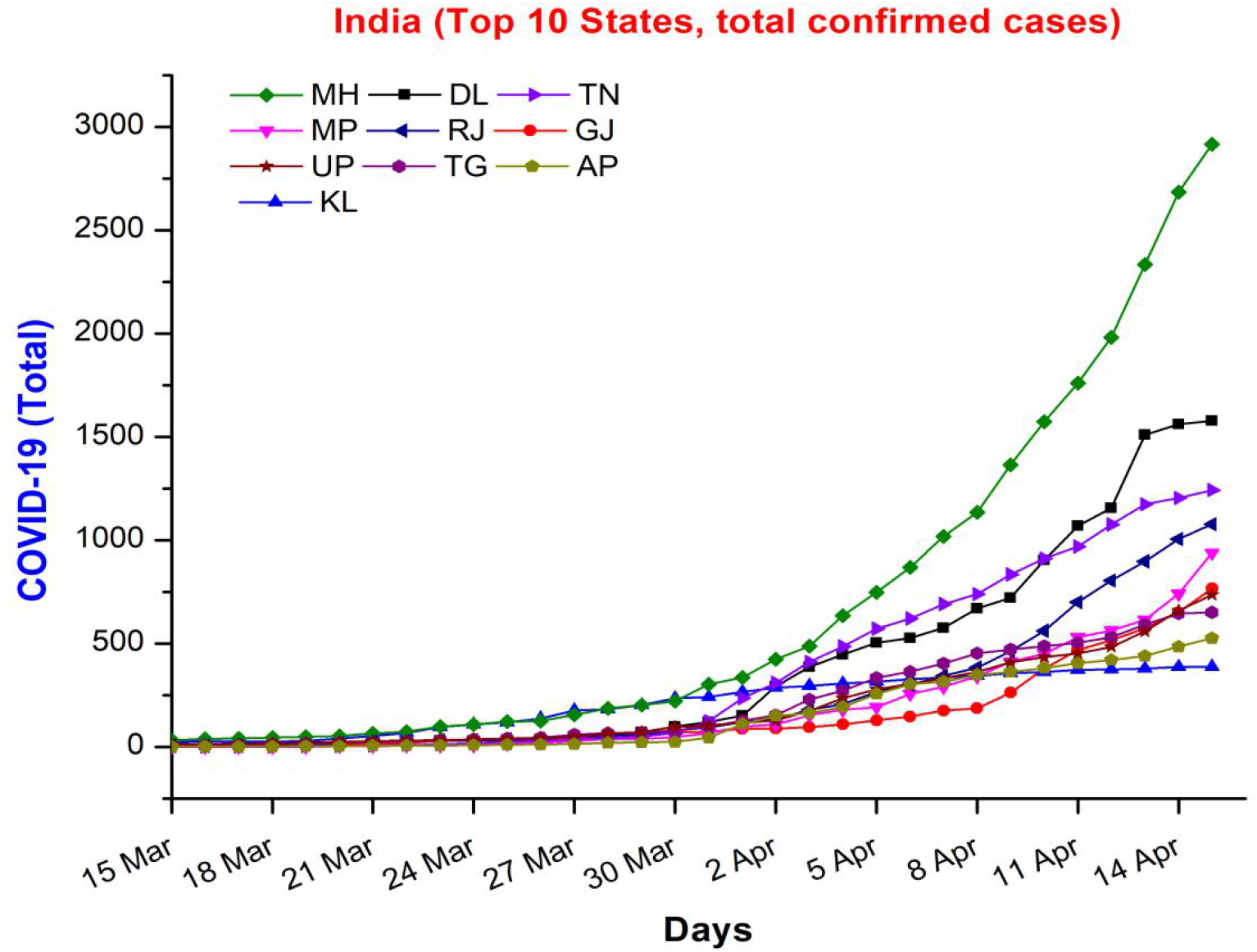
Top 10 states of India in total confirmed COVID-19 cases from 15 March-15 April 2020, different colors represent different states.

Since 30 January 2020, after the first case was reported in India, the increasing number of cases caused by COVID-19 had been identified until February. Up to the second week of March, only cases which were coming from foreign or in contact with them were reported, but after 15^th^ March, new daily cases appeared with no foreign travel cases. The number shoots up after 25^th^ March when a community of “Tablighi Jamat” in Delhi was reported that they might have COVID positive cases with at least 2000 people. After this event increasing number of daily cases continues with new and with those who have contact with these peoples. Presently this community has 30% cases of COVID-19 on total Indian cases across the India. On March 25, 2020, India, the residence of more than 1.38 billion humans, was forced to shut down both outbound and inbound traffic to contain the COVID-19 outbreak.

In addition to population mobility and human-to-human contact, environmental factors can impact droplet transmission and survival of viruses (e.g., influenza) but have not yet been examined for this novel pathogen for Indian cases. Absolute humidity, defined as the water content in ambient air, is a strong environmental determinant of other viral transmissions (Barreca and Shimshack, 2012; Luo et al., 2020). For example, influenza viruses survive longer on surfaces or in droplets in cold and dry air - increasing the likelihood of subsequent transmission. Thus, it is key to understand the effects of environmental factors on the ongoing outbreak to support decision-making about disease control, especially in locations where the risk of transmission may have been underestimated, such as in humid and warmer locations. We examine variability in Temperature (Tem), relative humidity (RH), and absolute humidity (AH) and transmission of COVID-19 across India. We show that the observed patterns of COVID-19 are not completely consistent with the hypothesis that high AH may limit the survival and transmission of this new virus.

Bu et al., (2020) found from a global perspective, cities with a mean temperature below 24 °C are all high-risk cities for 2019-nCoV transmission before June. In our case, it is not true as in India; the temperature was always high when the COVID-19 growth rate is high. Few studies supporting the hypothesis that high temp and high humidity will reduce the case like, Wang et al., (2020) find in their study, under a linear regression framework, high temperature and high humidity significantly reduces the transmission of COVID-19. They reported that a one-degree Celsius increase in temperature and a one percent increase in relative humidity lower *R* by 0.0225 and 0.0158, respectively. The transmission of coronaviruses can be affected by several factors, including climate conditions (such as temperature and humidity), population density, and medical care quality (Wang et al., 2020). Therefore, understanding the relationship between weather and the transmission of COVID-19 is the key to forecast the intensity and end time of this pandemic.

The number of 2019-nCoV cases detected in a country/state depends on multiple factors, including testing, population (density), community structure, social dynamics, governmental policies, global connectivity, air and surface life, reproduction number, and serial interval of the virus. Many of this information regarding 2019-nCoV are still emerging, such as the virus being airborne for more than 3 hours and having very different survival times on metals, cardboards and plastics (van Doremalen et al., 2020). The behavior of 2019-nCoV with meteorological parameters and with aerosols is still under investigation and also the subject of this paper over the Indian region during its spread and during the first lockdown period (25 March-15 April 2020). The analysis presented in this paper provides a direct comparison between the spread of COVID-19 virus and local environmental conditions over India region and study the growth rate of COVID-19 among different states of India (Fig.1).

## 2. Data and methodology

In our study, we have used COVID-19 data of daily conformed cases, surface temperature and surface humidity, aerosol optical depth (AOD), and NO_2_ with daily averages time series data for the Indian region. Following data were used,

### 2.1 Epidemiological data

The daily number of confirmed cases of patients infected with COVID-19 were taken from the WHO website and other public sources like Worldometer (Worldometer) and COVID-19Tracker/India (https://www.covid19india.org/) (COVID/Tracker) for March and April 2020.

### 2.2 Weather data

The meteorological parameters during the outbreak of the novel coronavirus in India for 2020 and three year past data were collected and analyzed. Air temperature and relative humidity data were taken from Atmospheric Infrared Sounder (AIRS) onboard EOS Aqua. Daily Data were taken for the surface temperature and surface relative humidity for the Indian region with resolution 1 degree for days March 1 to April 15, 2020, and for 2017-2019 from 1 March to 30 April. We have calculated the absolute humidity using these two parameters using the Clausius Clapeyron equation (Bolton, 1980) as follows:

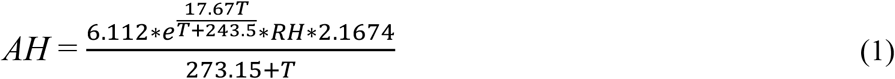

where AH is the absolute humidity, and T is the temperature in degrees C.

### 2.3 AOD and Pollutants data

Aerosols optical depth (AOD) and NO_2_ data were taken from GIOVANNI NASA (https://giovanni.gsfc.nasa.gov/giovanni/) sites from MODIS and OMI satellites. MODIS provides daily AOD data with the 1-degree resolution, which we used for the Indian region, and OMI also provides daily data with resolution 0.25 degrees. More details about MODIS data can be found elsewhere (Kumar et al., 2015). We have used data for 2017-2019 from March 1 to April 30 and 2020 from 1 March to 15 April.

### 2.4 HYSPLIT Trajectory data

HYSPLIT (NOAA) forward trajectories model was used to find out the surface air movement for the spread of aerosol.

## 3. Results and discussion

### 3.1 Variability of COVID-19 cases with the meteorological parameters (temperature, humidity, and absolute humidity)

We look into the relation between daily temperature (Tem), Relative humidity (RH), and absolute humidity (AH) with the daily number of confirmed cases of corona patients in India. We have used daily averaged meteorological data for the last three years (2017-2019) for March and April month and the same for the year 2020 for March 1 to April 15. We have plotted the average Tem of the last three years (2017-2019) and Tem of the year 2020 for the March and April months with the daily confirmed cases of COVID-19 in India. We found that average temp of the last three years during March and April was varying from 24 to 32 ^°^C, where this year Tem was underestimated from the last three years average Temp and varying from 22 to 30^°^C till 12 April. Both Temp showing continuously increasing trends for March and April. A maximum number of new confirmed cases appeared during the lockdown and showing an almost continuous increasing trend, and this shows there is a positive association of the new confirmed cases of COVID-19 with the increasing Tem of the region (Fig. 2).

**Fig 2.**
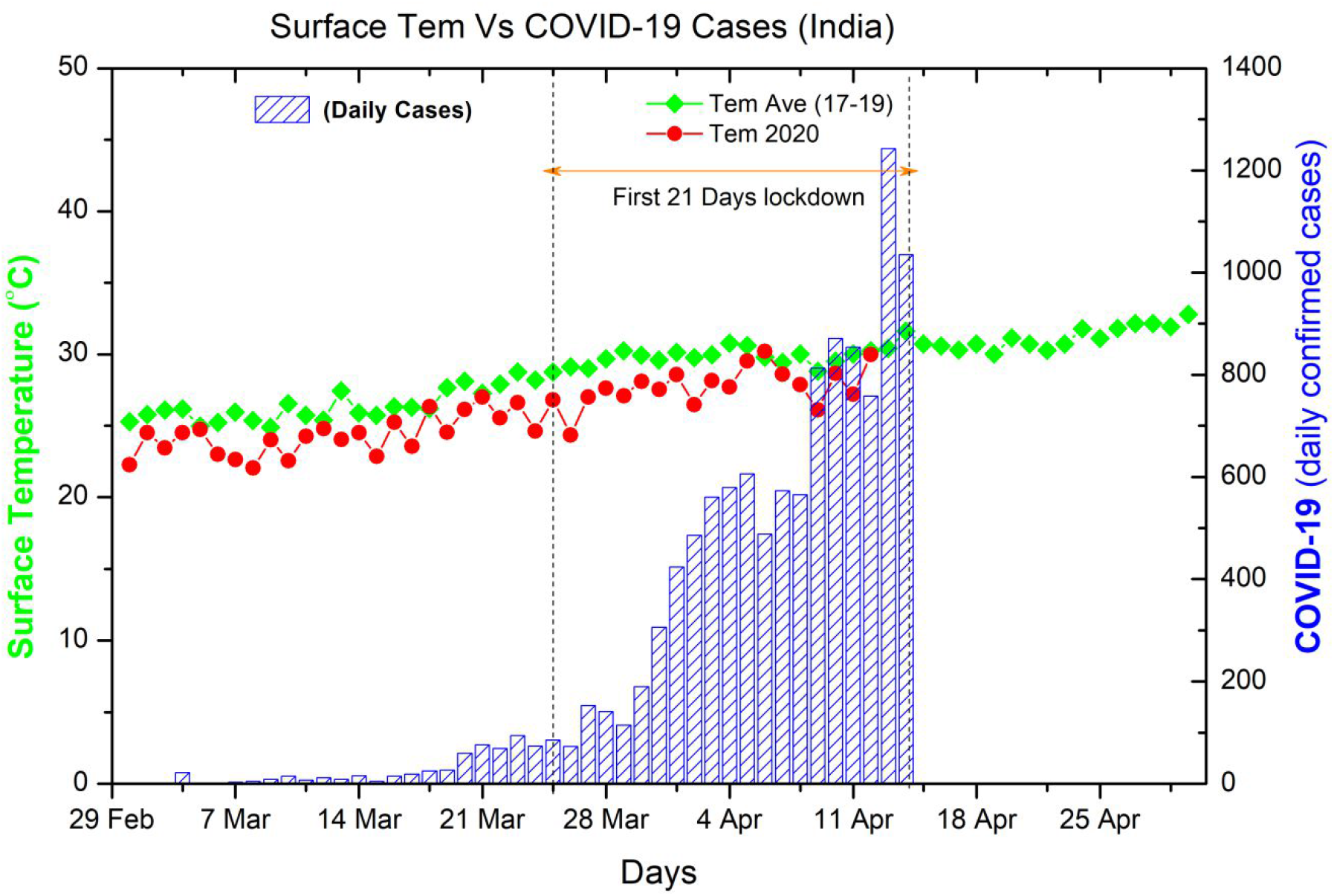
Diurnal surface temperatures Vs. Daily confirmed COVID-19 cases for the Indian region for March and April months for average Tem (2017-2019) and 2020.

Similarly, we have plotted the RH value and daily confirmed cases of COVID-19 for average (2017-2019) and for 2020 for March and April. We found a slowly decreasing trend in average RH varying from (42%-30%), and for 2020 we found overestimating RH from average with range 51% to 32% in India. RH values show the negative association with the daily confirm cases (Fig. 3). We found a positive correlation (Pearson’s r=0.56) between temperature and daily COVID-19 cases over India and found negative correlations of humidity (RH and AH) with daily COVID-19 Cases (Person’s r=-0.62, -0.37) (Graphical abstract).

**Fig 3.**
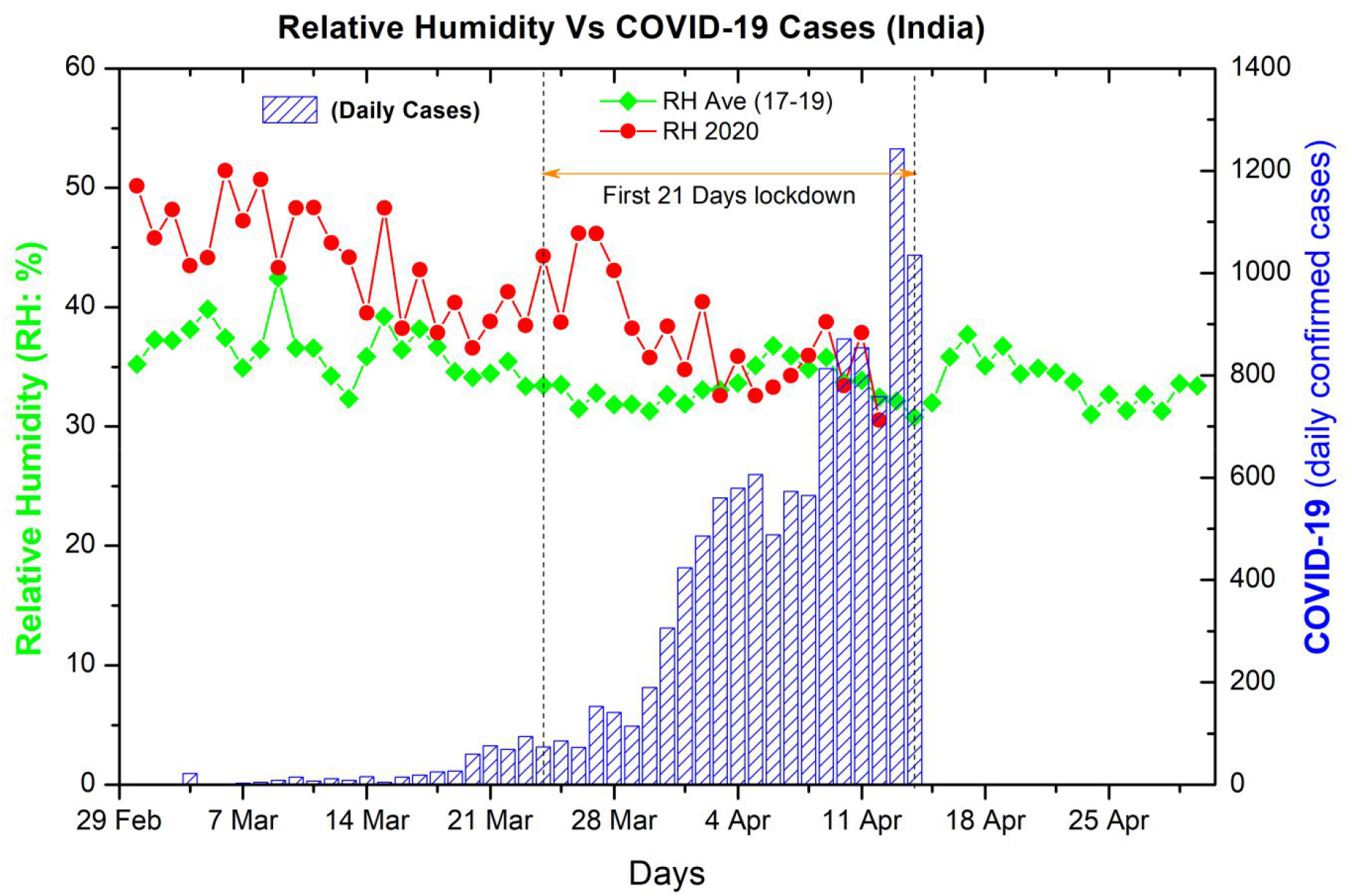
Same as Fig. 2 but for Relative humidity.

Absolute humidity, the mass of water vapor per cubic meter of air, relates to both temperature and relative humidity. We have also investigated the relation of confirmed cases with AH. The AH vales vary from 9 -11.5 g/m^3^ during March and April months of 2020. We found that the new cases are low when AH vales are greater than 9.5 g/m^3^ and high cases and high death when AH vales are going low from 9.5 g/m^3^ (Fig.4), this indicated the high values for AH might be helpful to reduce the new cases. Still, it’s not always true in Indian contest as the most affected states in India are Maharashtra and Tamil Nadu, which are the coastal states and have high values of Tem and RH through March to April. Our results show some similarities to the other studies done earlier on COVID-19 cases in China, Europe, and the USA.

Studying Wuhan, Guangzhou, and Beijing COVID-19 cased Bu et al., (2020) found a meteorological condition with the temperature between 13-19°C and humidity between 50% and 80% was suitable for the survival and transmission of the coronavirus. They also speculated that from February to the end of May 2020, the areas ranging the temperature (13 °C -24 °C) are all key areas for disease prevention, especially densely populated cities; from June and after that, the disease in regions with a mean temperature over 24 °C will begin to subside. They suggested before June 2020, cities with a global average temperature below 24 °C are all under high-risk of the transmission of new coronaviruses. After June, the risk of disease transmission will be significantly reduced in the cities with a mean temperature reaching 24 °C or higher. Our results for the Indian case did not match with the hypothesis said above as average temperature for the Indian region for March and April is above 24°C, even the COVID-19 cases are increasing rapidly in India even though restrict lockdown is there.

Based on their study of the spread of 2019-nCoV, Bukhari and Jameel (2020) hypothesize that the lower number of cases in tropical countries might be due to warm-humid conditions, under which the spread of the virus might be slower as has been observed for other viruses. They found that the relation between the number of 2019-nCoV cases and temperature and absolute humidity observed is strong; however, the underlying reasoning behind this relationship is still not clear. Similarly, they do not make clear that which environmental factor is more important. It could be that either temperature or absolute humidity is more important, or both may be equally or not important at all in the transmission of 2019-nCoV. The humidity dependency may be due to the less effective airborne nature of the viruses at higher absolute humidity, thus reducing the overall indirect transmission of 2019-nCoV at higher levels of humidity. Although higher humidity may increase the amount of virus deposited on surfaces, and virus survival time in droplets on surfaces, the reduction of the virus spread by indirect (through air) transmission may be the factor behind the reduced 2019-nCoV spread in the humid climate. These explanations are speculative and based on patterns observed for other coronaviruses. Urgent study/experiments on the association between coronavirus transmission against temperature and humidity in laboratories are needed to understand these associations.

### 3.2 Is COVID-19 airborne

Many studies suggested that COVID-19 may be stable up to 3 hours on aerosols (van Doremalen et al., 2020) and may be transmitted to long distances in a closed environment (Santarpia et al., 2020) as well as the open environment (Wang and Du, 2020). These studies suggest that COVID-19 may be airborne and can give a high risk of transmission through aerosols. Aerosols are particles formed by solid or liquid particles dispersed and suspended in the air. They contain soil particles, industrial dust particles, particulates emitted by automobiles, bacteria, microorganisms, plant spore powders, and other components. When a person who was infected with the virus, coughs, sneezes, breathes vigorously, or speaks loudly, the virus will be excreted from the body. It may dissolve with the aerosol and become the bio-aerosols. Bio-aerosols ranging in size from 1.0 to 5.0 μm generally remain in the air, whereas larger particles are deposited on surfaces. Droplets spread in the space of about 1 to 2 m from the source of infection. However, aerosol can travel hundreds of meters or more.

Wang and Du (2020), in his many case studies of COVID-19, speculated that the spread of the virus might be due to the aerosols because new patients in Inner Mongolia and Wuhan were never in direct contact with the confirmed cases. They found that COVID-19 may transmit through aerosol directly, but it needs to be further verified by experiments. If the aerosols can spread COVID-19, prevention and control will be much more difficult. Many of the information regarding 2019-nCoV are still emerging, such as the virus being airborne for more than 3 hours and having very different survival times on metals, cardboards and plastics (van Doremalen et al., 2020). Doremalen et al., (2020) found in his experiment that the COVID-19 virus can remain viable in aerosols throughout his experiment (3 hours), similar to that observed with SARS-CoV-1. Santarpia et al., (2020) in their clinical study found that SARS-CoV-2 is shed during respiration, toileting, and fomite contact, indicating that infection may occur in both direct and indirect (through aerosols) contact. Although this study did not employ any size-fractionation techniques to determine the size range of SARS-CoV-2 droplets and particles, the data was suggestive that viral aerosol particles are produced by individuals that have the COVID-19 disease, even in the absence of cough. Therefore the COVID-19 may spread through the aerosols if the sufficient amount of aerosols is present in the environment. Also, the reduction in aerosol concentration during the lockdown may reduce the risk of transmission of COVID-19 through aerosols.

### 3.3 Association of COVID-19 with aerosols and pollutants in India

#### 3.3.1 Association of COVID-19 with AOD

To prevent further spread of COVID-19, India has started the world’s biggest lockdown of history on 25^th^ March 2020, where the whole country was locked, and more than 1.38 billion people were forced to remain in their homes. This lockdown surely has positive feedback by less daily new COVID cases compared to Europe and the USA and also affect the environment. Due to the strict lockdown in India, all public transport, industries, and individual activities were shut, which was reflected in air quality and aerosols over India. The aerosols decrease sharply over India in comparison with the average value of AOD of the last three years (Fig. 5). We have found a clear decrease in AOD value from the first day of lockdown with little increase in value in the coming days due to some relaxation in lockdown. Also, April month is harvest month so, aerosols are increased due to harvesting and crop residue burning in the April month, which can also be seen in averaged data. Now the first 21 days lockdown (25 March-15April) period is over, and the second lockdown (15 April-3 May) is going on but with some relaxation in few important activities, so It may increase the aerosol concentration. If a lockdown is followed strictly till May 3, 2020, then a big reduction in AOD may be observed, which may restrict the risk of further new COVID-19 cases by its transmission through aerosols.

**Fig 4.**
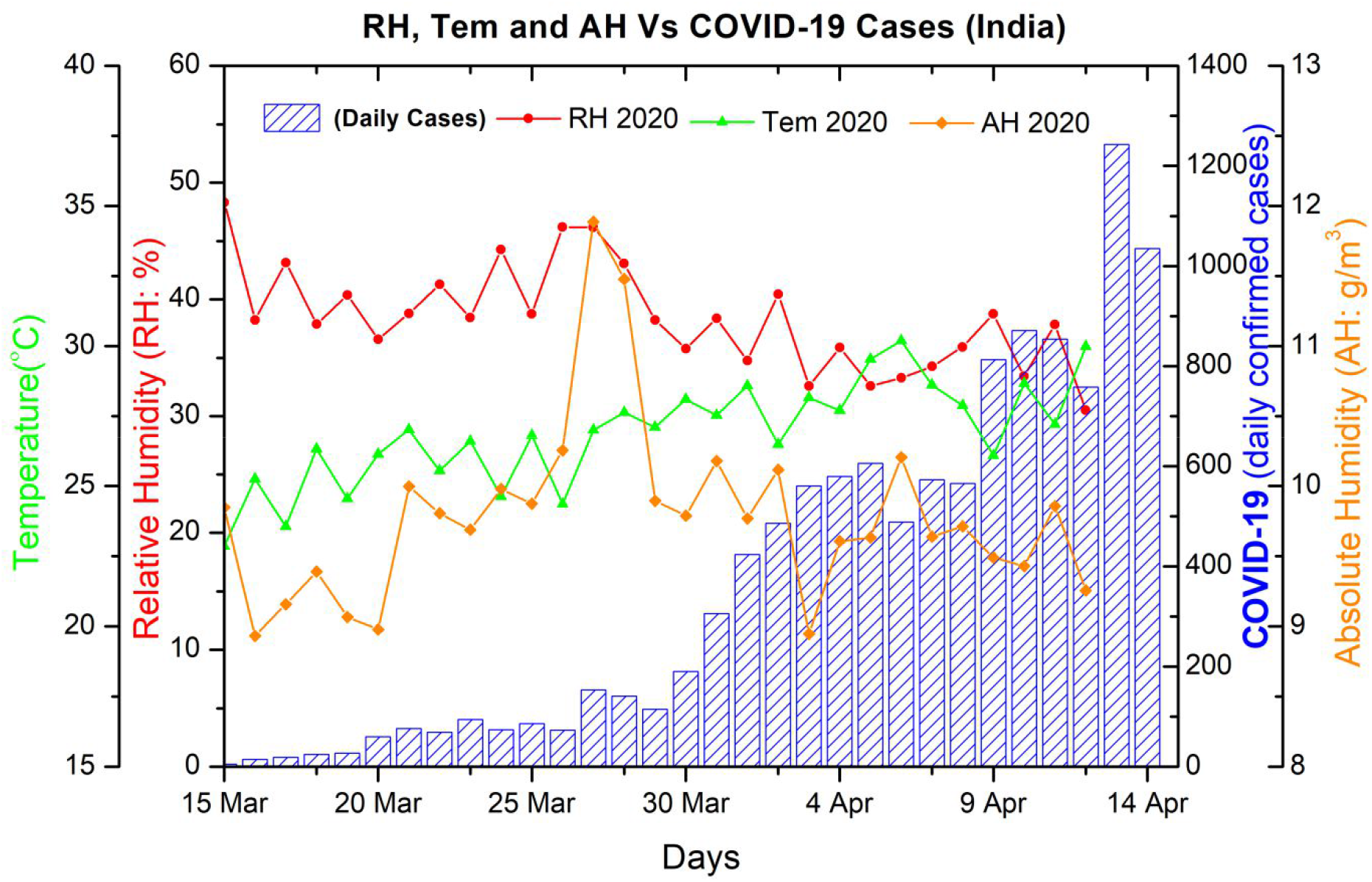
Diurnal variations of surface temperatures, relative humidity, and absolute humidity Vs. Daily confirmed COVID-19 cases for the Indian region for 15 March-15 April 2020.

**Fig 5.**
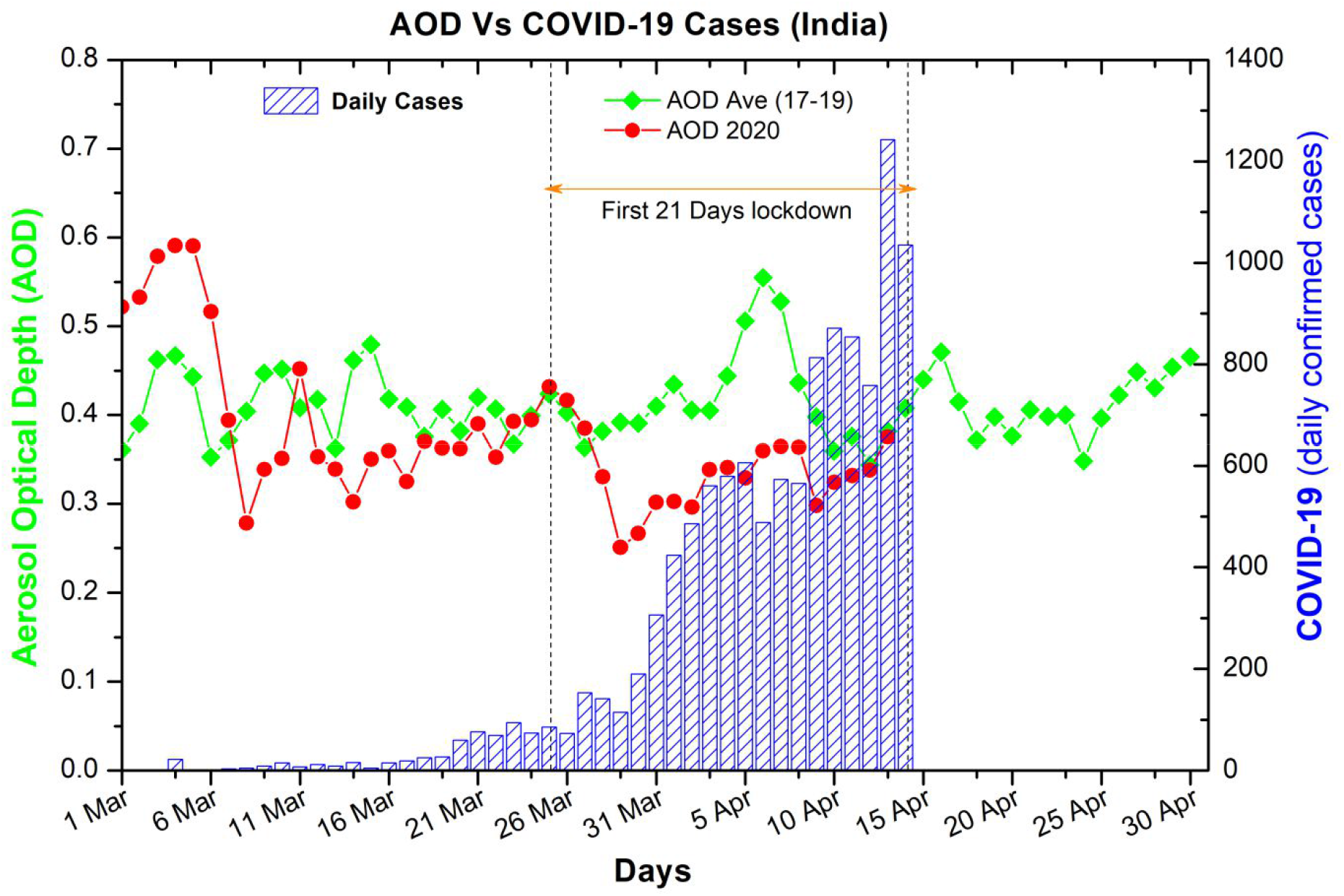
Diurnal variations of AOD Vs. Daily confirmed COVID-19 cases for the Indian region for March and April months for average AOD (2017-2019) and AOD for 2020.

#### 3.3.2 Association of COVID-19 with other pollutants like NO_2_

NO_2_ is a marker of the particulate matter (PM 2.5) for the study of pollutions. Strict lockdown in India reduces the tropospheric column NO_2_ also, which again lowers the risk of COVID transmission through PM 2.5 for the Indian region (Fig. 6). It’s very clear from the plot that the concentration of NO_2_ is reduced after the lockdown in comparison to the three-year average value of NO_2_.

**Fig 6.**
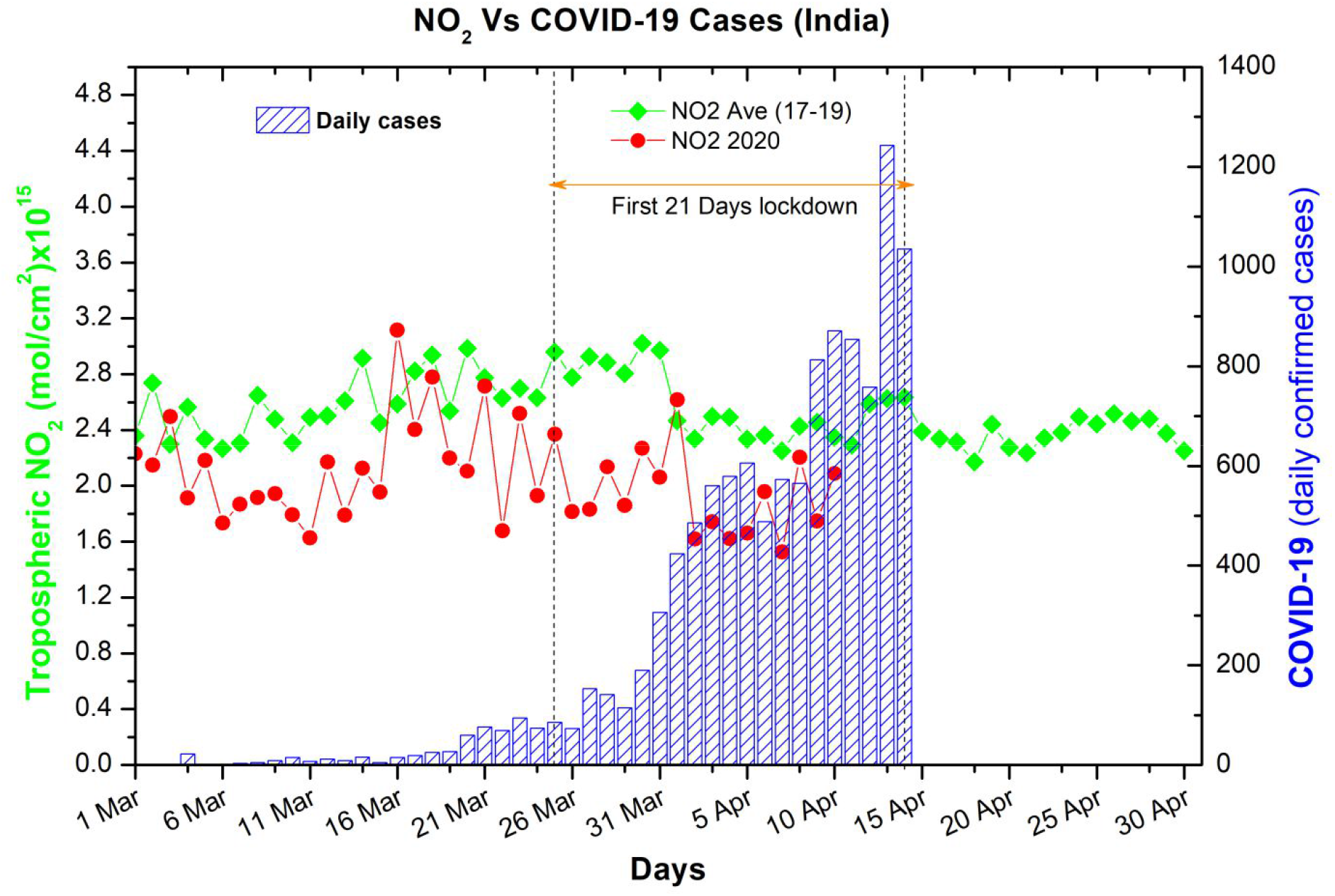
Same as Fig.5 but for diurnal NO_2_.

#### 3.3.3 Percentage dropdown in pollutants concentration during the first lockdown

Figure 7 shows the percentage dropdown in the values of AOD and NO_2_ in comparison to the average value of the last three years over India. We found maximum dropdown during the lockdown period of 2020 up to 36% for AOD and 37 % for NO_2_ in comparison to the average value of 2017-2019.

**Fig 7.**
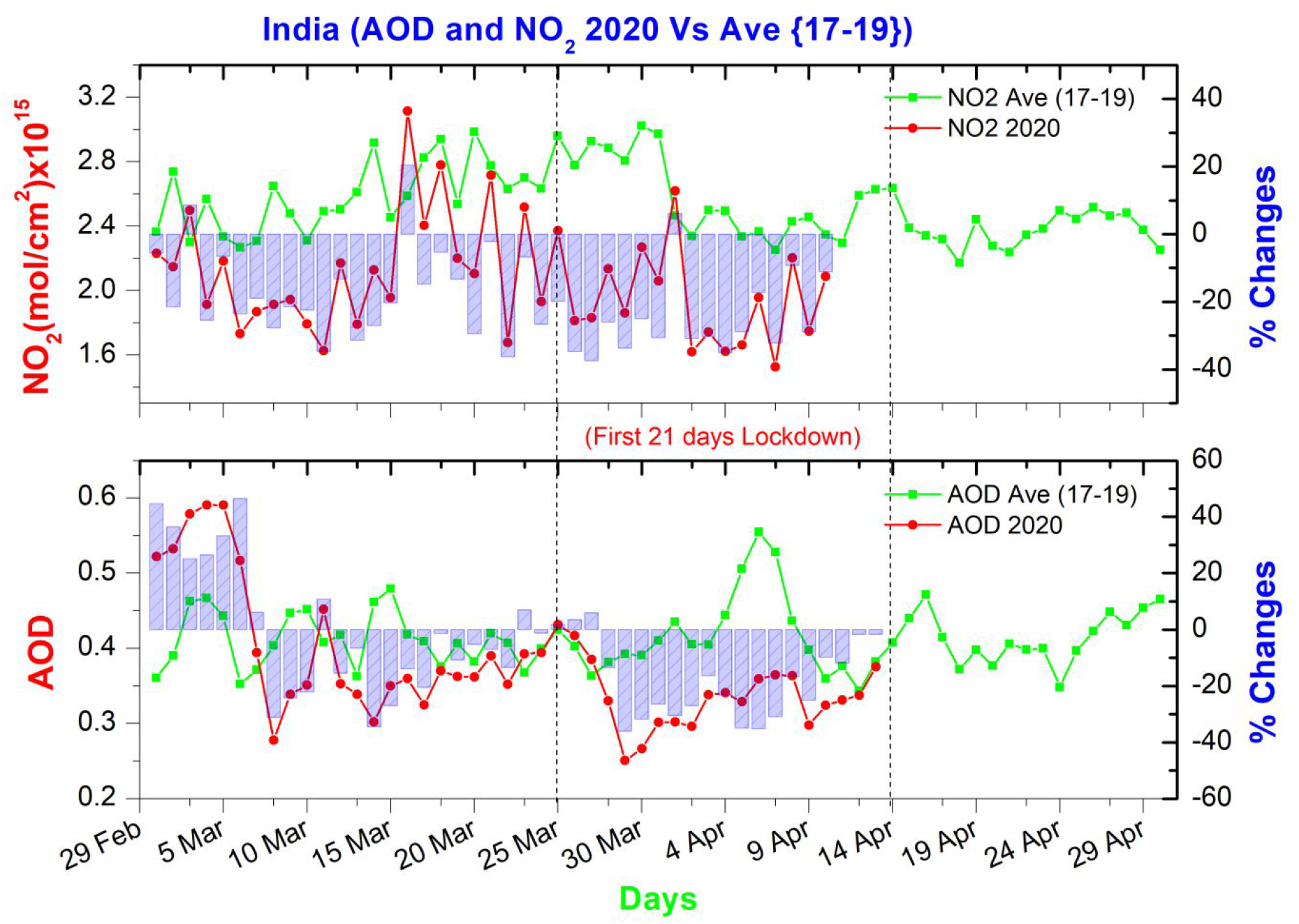
Percentage changes (Column bar) in diurnal variations of AOD and NO_2_ for the Indian region for March and April months for average (2017-2019), and for 2020, the first lockdown period is between two dashed vertical lines.

#### 3.3.4 Correlation of AOD with COVID-19 cases

We have also investigated the correlation of AOD with the daily new confirmed case in India for March and April. We found the negative correlation during March when there were fewer cases of COVID-19, but in April, the correlation turned in to positive (Fig. 8) when daily cases are more and which again indicating that possible COVID-19 transmission through the aerosols if it is airborne.

**Fig 8.**
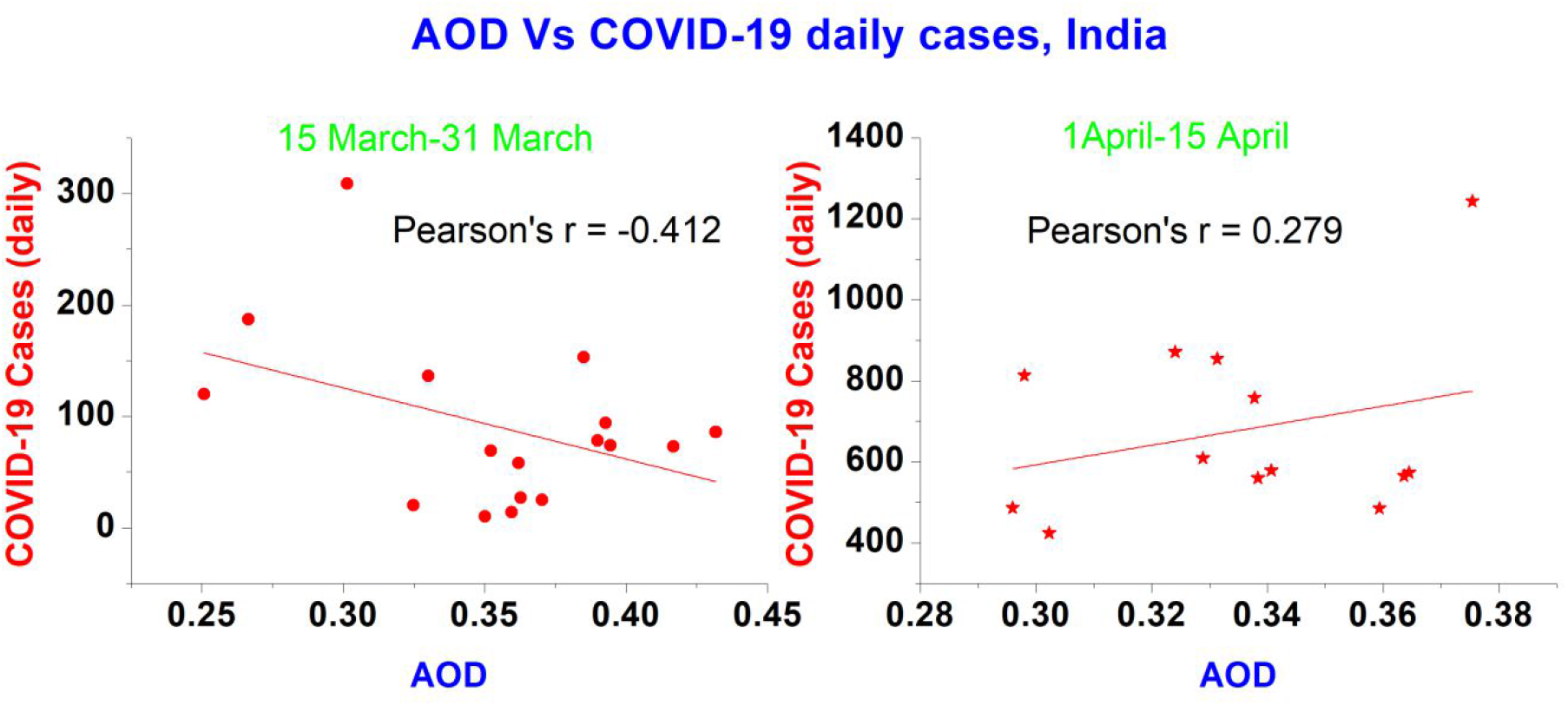
Correlation of daily AOD Vs. daily confirmed COVID-19 cases for Indian region for 15 March -15 April for 2020.

### 3.4 Trajectory analysis of aerosols emitted at surface level

We have used the HYSPLIT trajectory model for calculation of the forward trajectory (not shown here) of surface aerosols. As van Doremalen et al., (2020) found in his study that COVID-19 virus can be stable at aerosols surface for about 3 hours. So if a coronavirus is attached to the aerosols, then it may travel for longer distances and become airborne, this may be a reason for the high number of cases in the USA and other European countries as in early-stage they did consider COVID-19 may not transmit through the air and not using the face mask. So when we run HYSPLIT forward trajectory model for the surface level air for 3 hours period, we found that in normal condition in April month in India, a surface level aerosol can travel up to 4 km distance in 3 hours according to the wind speed and direction. So there is a great risk of transmission of COVD-19 through aerosols if their concentration is high and viruses have plenty of bases to stick it. Therefore our study suggests that there must be strict lockdown for all factors affecting the concentration of aerosols; otherwise, it may be an invitation to a disaster to give relax in lockdown in India like a country with high aerosols in coming months.

### 3.5 Implications for preventing future transmission of 2019-nCoV

Before in March 2020, many studies speculated that the places with higher temperatures are in less risk, and it appeared that temperature might play an important role in the spread of the virus. However, more new cases were recorded in regions with a temperature between 16 and 18^°^C in March even more up to 30^°^C in India during March and April 2020, which is now challenging the hypothesis that a rise in temperature would minimize the spread of the 2019-nCoV. Nonetheless, the observation of more than 10,000 cases in the last ten days in India makes it clear that the effect of rising summer temperature, if any, would not be observed in the current hotspot of India, as the mean temperature for most of the major cities in India is above 30^°^C for most of April and May. Indeed, laboratory experiments performed between 21-23^°^C at a relative humidity of 40%, showed that the virus survived for several days on plastics and metals (van Doremalen et al., 2020). Therefore temperature may affect similar to the relationship observed between SARS-CoV and temperature (Bukhari and Jameel, 2020). Still, we can’t say at what temperature and to what extent it would help in reducing the spread of 2019-nCoV. Under any circumstances, we believe that large gatherings (both indoor and outdoor) should be avoided across India.

Based on currently available data, 2019-nCoV is spreading easily in regions with absolute humidity <10 g/m^3^ (Bukhari and Jameel, 2020), this has serious implications on the assumption that the 2019-nCoV spread would slow down during summer in the current hotspot of India as in many regions the absolute humidity might be above 10 g/m^3^ (Fig.4). On the other hand, if, new cases in April and May continue to cluster within the current observed range of AH, i.e., 9 to 11 g/m^3^, then the states experiencing monsoon having a high absolute humidity (>11 g/m 3) may see a slowdown in transmissions, due to climatic factors. If it is then for India, it has long waiting time up to June 15, when the monsoon will come, and up to then, only government mitigation policies are going to reduce the COVID-19 spread over India.

## 4. Conclusions

The novel coronavirus pneumonia is caused by 2019-nCoV, which is a new pathogen for the human being. Due to its outbreak during the spring and summer, it is spreading very fast in a short period. Faced with this new disease, we lack reliable epidemiological information for effective treatment and prevention. However, any infectious disease origins and spread occur only when affected by certain natural and social factors through acting on the source of infection, the mode of transmission, and the susceptibility of the population. The weather and meteorological factors also may play a part in the coronavirus outbreak besides the social factors. Also, aerosols may play a crucial role during the spread of COVID-19. Following conclusions and suggestion may be drawn from the current study;

1. We have studied the effect of environmental factors and aerosols on the spread of COVID-19 during March and April in India. We have studies the total number of daily confirmed cases of COVID-19 and its association with the temperature, relative humidity, and absolute humidity over India for March and April 2020. We found a positive association between daily new cases of COVID-19 with temperature for India. Relative humidity and Absolute humidity is negatively associated with daily cases of COVID-19 in India. We found a positive correlation (Pearson’s r =0.56) between temperature and daily COVID-19 cases over India. We found a negative correlation of humidity (RH and AH) with daily COVID-19 Cases (Person’s r =-0.62, -0.37).
2. To reducing the COVID-19 effect, summer alone is not going to help majorly, as high absolute humidity regions like Maharashtra and Tamil Nadu are getting the maximum number of confirmed daily COVID-19 cases (Fig. 1). So only the government’s strict mitigation strategies would be helpful, whether their totals lockdown or strictly imposing the social distancing for the community. Two coastal states of India (Kerala and Maharashtra) are the example for rest of the India who has an almost same meteorological environment and had same numbers of COVID-19 patient on 29^th^ March, but during first lockdown period, their governments’ strategies made a big difference as now Kerala is on 11^th^ position, and Maharashtra is at the top in the total number of COVID-19 patient in India (Fig.1).
3. There is a thin hope that increasing temperature in coming months in India may reduce the number of new COVID-19 cases, because in India during summer, crop residue burning and major dust storms occur (Kumar et al., 2015), which further will degrade the air quality especially in the north Indian region and will create the health emergency by producing respiratory diesis due to increased particulate matter, ultimately which may enhance the number of death of patient suffering from the COVID-19, also will increase the chance of spreading through aerosols.
4. We also investigated the aerosols and pollutants behavior with the total number of cases during the COVID-19 outbreak. The concentration in aerosols (AOD) and other pollutants (NO_2_) was sharply reduced during the first lockdown period (25 March-15 April) in India, which may have a negative effect and lowered the risk of COVID-19 to be airborne because less available aerosols as the base for the virus to stick on it.
5. Since many earlier studies and our findings are suggesting that COVID-19 may be airborne, so to slow down its spread, governments should impose strict lockdown and motivate the society to follow the social distancing, thermal scanning, and wearing a face mask when going outside. Health workers and those are working at the front line must take precautions as it may be airborne. Also, the mild infected and the severely infected patient should be kept in a separate medical facility.
6. The structures of social contact critically determine the spread of the infection and, in the absence of vaccines, the control of these structures through large-scale social distancing measures appears to be the most effective means of mitigation. As in the previous study by Singh and Adhikari, (2020) suggested that the three-week lockdown will be insufficient, and it’s found true for India, our suggestion is for complete lockdown with minimal relaxation with strict social distancing for 4 more weeks.
7. We found that early lockdown in India reduces the possible number of infections/death due to the coronavirus. In India, most of the cases identified during the lockdown period, which shows less effect of weather to slow it down, as it is already summer in the Indian region. The number of cases per million populations is least for India in comparison to the USA and Europe, which shows the government mitigations working well as it was a historic move by the Indian government to lockdown 1.38 billion people with the different social and economic background when there were only 600 cases of the corona.
8. After April, the Temperature will be very high as summer approaching its maximum and in India during June also humidity will be high in coastal states. It will increase slowly over the northern parts of India in late June month. Then this hypothesis may be tested for the Indian region that high Temp and high humidity will reduce the number of cases due to the coronavirus. That time will tell, but before that, at least up to 15 May 2020, there must be strict lockdown by the government (the second lockdown announced for 15 April-3 May) over India and community should strictly follow the social distancing to reduce the number of COVID-19 cases. Relaxing in the lockdown and environmental rules in terms of pollutant emissions from power plants, factories, and other facilities would be the wrong choice. It could increase pollutants like PM2.5, which may result in more COVID-19 incidences and deaths in India. There is a need for a more appropriate study of the rate of outdoor transmission versus indoor and direct versus indirect transmission as they are not well understood, and environmental-related impacts are mostly applicable to outdoor transmissions.

## Data Availability

All data are available on the internet through websites.

## Acknowledgment

We are thankful to GIOVANNI NASA for providing the satellite data for AOD, NO_2_, Temperature, and Relative humidity. We are thankful to the volunteers of COVID-19 Tracker/India (https://www.covid19india.org/) for providing all the data of India related to COVID-19. We thank NOAA for providing the HYSPLIT trajectories through their online models. Finally, the author is profoundly grateful to the Vice-Chancellor of V.B.S. Purvanchal University Prof. (Dr.) Raja Ram Yadav for providing funding and encouraging to engage in research with teaching.

